# Likelihood of being Physically Inactive from a Nationally Representative sample of Autistic Children

**DOI:** 10.1101/2021.11.05.21265973

**Authors:** Vijay Vasudevan

## Abstract

Despite the many health risks of physical inactivity, studies have demonstrated individual, family, and environmental determinants of inactivity for autistic children. However, these studies never examined these correlates at the same time. Therefore, the purpose of this study was to explore these ecological domains concurrently when examining physical inactivity correlates for autistic children. This study used data from the 2016-2020 National Survey of Children’s Health. The authors predicted physical inactivity while controlling for child, parental/household, and neighborhood correlates with autism status as the comparison group. When controlling for covariates, children with co-occurring autism and intellectual and developmental disability (IDD) (adjusted odds ratio (aOR)= 1.91, 95% confidence interval (CI): 1.36-2.68) or ASD only (aOR = 1.91, CI: 1.48-2.48) were significantly more likely to be inactive when compared to children without autism or IDD. However, autism medicine and autism severity were not predictors for obese autistic children. These findings indicate that it is important to take a holistic, ecological approach when exploring the correlates of inactivity for autistic children.

## INTRODUCTION

The health risks of being overweight and obese are well known including: type 2 diabetes, high blood pressure, coronary heart disease, sleep apnea and breathing problems, depression, anxiety, and body pain.^1^ Autistic children and adolescents are at greater risk of being overweight/obese and having unmet healthcare needs.^2,3^ Some of the contributing factors to being obese for the autistic children include physical inactivity, sleep quality, and parental concern about weight.^4-6^ McCoy (2020) found an inverse relationship between being active seven days a week and obesity status of autistic children.^7^

Physical activity has been shown to improve weight status while also addressing the health risks of being obese, such as improved cardiorespiratory health and muscular fitness, improved cardiometabolic health, and reduced risk of depression.^8^ To reach these benefits, all children and youth are recommended to get at least 60 minutes of moderate to vigorous physical activity every day.^8^ Despite these benefits, approximately a quarter of all children age six to 17 met this guideline.^9^ Children and adolescents with disabilities had significantly lower odds of meeting the physical activity guidelines.^10^ Barriers to physical activity for people with disabilities exist at multiple domains of the ecological model, including individual factors like health, family perception, and having a supportive environment.^11,12^

A previous study, by Case (2020) which explored the likelihood of meeting the physical activity guidelines, found that children with an autism diagnosis were 35% less likely to meet the physical activity guidelines when compared to children without an autism diagnosis. However, in an adjusted model for autism severity, having multiple developmental disabilities, and having a health condition that limits the child’s ability to do activities, autism was not a significant predictor of meeting the physical activity guideline.^13^ This study did not control for important covariates such as sleep, obesity status, parental concern about weight, and living in a supportive neighborhood. Therefore, the purpose of this study was to explore the likelihood to meet the physical activity guidelines by autism status while controlling for ecological correlates.

## METHODS

### Design

This study used combined data from the 2016-2020 National Survey of Children’s Health (NSCH). The NSCH is a nationally, representative sample survey which is administered annually by the US Census Bureau for the Maternal Child Health Bureau, Health Resources and Services Administration. The NSCH asks age-based topical questions (0-5 years, 6-11 years, and 12-17 years old). All responses are parent-reported. Individual survey weights were divided by the number combined survey years in order to derive nationally, representative estimates. Because NSCH data were publicly available, the authors did not seek Institutional Review Board approval.

### Autism Status

Because a third of all children with ASD had an intellectual disability (IDD),^14^ one mutually exclusive variable for autism status was created. which had the following responses: no autism and IDD, autism only, and autism with co-occurring IDD. The NSCH asks two questions to assess current status of ASD and IDD. Current autism status was determined by two questions: (1) “Has a doctor or other health care provider EVER told you that this child has Autism or Autism Spectrum Disorder (ASD)? *Include diagnoses of Asperger’s Disorder or Pervasive Developmental Disorder (PDD)*.” and (2) “If yes, does this child CURRENTLY have the condition?” Children whose parents who reported “yes” to both items were categorized with being “Currently diagnosed with autism.” If the parent reported “no” to either question, the child was categorized as “Not currently diagnosed with autism.” Current IDD status was determined by two questions: (1) “Has a doctor, other health care provider, or educator EVER told you that this child has… Intellectual Disability (formerly known as Mental Retardation)? *Examples of educators are teachers and school nurses*.” and (2) “If yes, does this child CURRENTLY have the condition?” Children whose parents who reported “yes” to both items were categorized with being “Currently diagnosed with IDD.” If the parent reported “no” to either question, the child was categorized as “Not currently diagnosed with IDD.”

Parents who reported that their child are currently diagnosed with ASD were also asked for their child’s autism severity (rated as mild, moderate, or severe) and if the child is taking medicine for autism.

### Physical Activity

Physical activity was asked of children who were from six to 17 years old. Physical activity was asked by the NSCH with one item: “DURING THE PAST WEEK, on how many days did this child exercise, play a sport, or participate in physical activity for at least 60 minutes?” Response options included: 0 days, 1-3 days, 4-6 days, and every day. The authors collapsed the responses into three categories: Inactive – 0 days, Insufficiently active – 1 to 6 days, and Active – 7 days. A second dichotomous variable was created: Inactive versus Active or Insufficiently Active.

### Correlates

The authors controlled for child, parent/household, and community variables. Child variables include age (6-11 years old and 12-17 years old), gender, race (White, non-Hispanic, Black, non-Hispanic, Hispanic, and Multirace or Other, non-Hispanic), obesity category (underweight, normal weight, overweight, and obese), and sleep. Parent/household variables include parent’s highest education, parent concern about child’s weight, and federal poverty level (FPL). The neighborhood variable was whether or not the child lives in a supportive neighborhood.

### Analyses

Stata version 16.1 was used for all analyses.^15^ Weighted, demographic prevalence estimates (%) and 95% confidence intervals (CIs) were calculated for survey respondents of all respondents whose child was at least six years old. Chi-square analysis was performed to identify differences in physical activity level (inactive, insufficiently active, and active) and autism status. Unadjusted and adjusted odds ratios (aOR) were calculated, using logistic regression, to predict physical inactivity (inactive versus active or insufficiently active) controlling for child, parental/household, and neighborhood correlates with autism status as the comparison group.

A separate logistic regression, of just autistic children who were obese, was performed to predict physical inactivity, while controlling for the same correlates stated above, and adding autism severity and whether or not the autistic child is taking medicine to treat their autism.

## RESULTS

Table 1 presents the prevalence estimates and CIs for the demographic variables of all survey participants, whose children were at least six years old. Approximately three percent of children were currently diagnosed with ASD. Roughly a third (34.8%) of parents reported that their children did not get the recommended hours of sleep. About 15% of parents reported that their children were overweight or obese (15.6% and 15.8% respectively), and 7.6% was concerned that their child’s weight was too high. Finally, 10.1% of parents reported that their child got zero days of at least 60 minutes of activity, while just 22.1% were active every day.

**Table 1:** Demographics.

|  | Prevalence Estimate | 95% CI |
| --- | --- | --- |
| Current Autism Status |  |  |
| Not currently diagnosed with autism | 96.9% | (96.7%, 97.1%) |
| Currently have autism | 3.1% | (2.9%, 3.3%) |
| Child age |  |  |
| 6-11 years old | 15.6% | (15.2%, 16.1%) |
| 12-17 years old | 84.4% | (84.0%, 84.9%) |
| Gender |  |  |
| Female | 48.9% | (48.4%, 49.4%) |
| Male | 51.1% | (50.6%, 51.7%) |
| Race |  |  |
| White, non-Hispanic | 50.7% | (50.2%, 51.3%) |
| Black, non-Hispanic | 13.2% | (12.8%, 13.6%) |
| Hispanic | 25.2% | (24.6%, 25.8%) |
| Multirace or Other, non-Hispanic | 10.9% | (10.6%, 11.2%) |
| Adequate amount of age |  |  |
| Child sleeps recommended age-appropriate hours | 65.3% | (64.7%, 65.8%) |
| Child sleep less than recommended age-appropriate hours | 34.8% | (34.2%, 35.3%) |
| Parent highest education |  |  |
| College degree or higher | 49.6% | (49.0%, 50.1%) |
| Some college or associate degree | 21.8% | (21.4%, 22.2%) |
| High school graduate or less | 28.6% | (28.1%, 29.2%) |
| Federal Poverty Level |  |  |
| Greater than or equal to 200% FPL | 58.6% | (58.0%, 59.1%) |
| Less than 200% FPL | 41.4% | (40.9%, 42.0%) |
| Does child live in a supportive neighborhood |  |  |
| Do not live-in supportive neighborhood | 45.5% | (44.9%, 44.6%) |
| Live in supportive neighborhood | 54.5% | (54.0%, 55.1%) |
| Parent-reported Child Weight Category |  |  |
| Underweight (less than 5th percentile) | 6.5% | (6.1%, 6.9%) |
| Normal weight (5th to 84th percentile) | 62.1% | (61.3%, 62.9%) |
| Overweight (85th to 94th percentile) | 15.6% | (15.0%, 16.2%) |
| Obese (95th percentile or above) | 15.8% | (15.2%, 16.5%) |
| Parent concerned about child's weight |  |  |
| No, not concerned | 89.4% | (89.1%, 89.8%) |
| Yes, concerned it is too high | 7.6% | (7.4%, 8.0%) |
| Yes, concerned it is too low | 2.9% | (2.8%, 3.1%) |
| Physical activity at least 60 minutes per day |  |  |
| Inactive - 0 days | 10.1% | (9.7%, 10.5%) |
| Insufficiently active - 1-6 days | 67.8% | (67.2%, 68.4%) |
| Active - 7 days | 22.1% | (21.6%, 22.7%) |

Children with co-occurring ASD and IDD or ASD only were statistically more likely to be “Inactive – 0 days” when compared to children who do not have ASD (21.0% and 17.3% versus 9.4% respectively, χ^2^=16.4, p < 0.001). When controlling for covariates (Table 2), children with co-occurring ASD and IDD (aOR = 1.91, CI: 1.36-2.68) or ASD only (aOR = 1.91, CI: 1.48-2.48) were significantly more likely to be inactive when compared to children without ASD or IDD.

**Table 2:** Likelihood of Being Inactive.

|  | Unadjusted Odds Ratio<br>Prevalence |  | Adjusted Odds Ratio<br>Prevalence |  |
| --- | --- | --- | --- | --- |
|  | Estimate | 95% CI | Estimate | 95% CI |
| Current Autism Status |  |  |  |  |
| Not currently diagnosed with autism<br>or intellectual/developmental disability | REF |  | REF |  |
| **Currently have autism only** | **2.01** | **(1.49, 2.70)** | **1.91** | **(1.36, 2.68)** |
| **Currently have autism and IDD** | **2.56** | **(1.88, 3.48)** | **1.91** | **(1.48, 2.48)** |
| Child age |  |  |  |  |
| 6-11 years old |  |  | REF |  |
| **12-17 years old** |  |  | **2.27** | **(1.73, 1.68)** |
| Gender |  |  |  |  |
| Male |  |  | REF |  |
| **Female** |  |  | **1.51** | **(1.35, 1.68)** |
| Race |  |  |  |  |
| White, non-Hispanic |  |  | REF |  |
| **Black, non-Hispanic** |  |  | **1.52** | **(1.31, 1.76)** |
| Hispanic |  |  | 1.04 | (0.90, 1.20) |
| **Multirace or Other, non-Hispanic** |  |  | **1.32** | **(1.12, 1.54)** |
| Parent highest education |  |  |  |  |
| College degree or higher |  |  | REF |  |
| **Some college or associate degree** |  |  | **1.27** | **(1.13, 1.43)** |
| **High school graduate or less** |  |  | **1.61** | **(1.40, 1.84)** |
| Federal Poverty Level |  |  |  |  |
| Greater than or equal to 200% FPL |  |  | REF |  |
| Less than 200% FPL |  |  | 1.07 | (0.94, 1.21) |
| Does child live in a supportive neighborhood |  |  |  |  |
| Live in supportive neighborhood |  |  | REF |  |
| **Do not live in supportive neighborhood** |  |  | **1.52** | **(1.36, 1.70)** |
| Parent-reported Child Weight Category |  |  |  |  |
| Normal weight (5th to 84th percentile) |  |  | REF |  |
| Underweight (less than 5th percentile) |  |  | 1.23 | (0.95, 1.59) |
| Overweight (85th to 94th percentile) |  |  | 1.08 | (0.94, 1.26) |
| **Obese (95th percentile or above)** |  |  | **1.21** | **(1.01, 1.47)** |
| Parent concerned about child's weight |  |  |  |  |
| No, not concerned |  |  | REF |  |
| **Yes, concerned it is too high** |  |  | **1.89** | **(1.59, 2.25)** |
| **Yes, concerned it is too low** |  |  | **1.67** | **(1.29, 2.18)** |
| Adequate amount of age-appropriate sleep |  |  |  |  |
| Child sleeps recommended age-appropriate hours |  |  | REF |  |
| **Child sleep less than recommended age-appropriate hours** |  |  | **1.16** | **(1.04, 1.30)** |

When exploring autistic children who were obese (Table 3), the significant correlates of physical inactivity were parent concern that their child’s weight is too high (aOR = 02.97, CI: 1.28-6.87) and the child’s age is between 12 and 17 years old (aOR = 5.18, CI: 1.63-16.51).

**Table 2:** Likelihood of Being Inactive for Autistic Children who are Obese.

|  | Adjusted Odds Ratio |  |
| --- | --- | --- |
|  | Prevalence Estimate | 95% CI |
| Child age |  |  |
| 6-11 years old | REF |  |
| **12-17 years old** | **5.18** | **(1.63, 16.51)** |
| Gender |  |  |
| Male | REF |  |
| **Female** | **0.40** | **(0.17, 0.93)** |
| Race |  |  |
| White, non-Hispanic | REF |  |
| **Black, non-Hispanic** | **0.13** | **(0.03, 0.49)** |
| Hispanic | 1.32 | (0.49, 3.60) |
| Multirace or Other, non-Hispanic | 0.76 | (0.24, 2.43) |
| Parent highest education |  |  |
| College degree or higher | REF |  |
| **Some college or associate degree** | **0.46** | **(0.21, 0.99)** |
| High school graduate or less | 0.46 | (0.19, 1.09) |
| Federal Poverty Level |  |  |
| Greater than or equal to 200% FPL | REF |  |
| Less than 200% FPL | 1.12 | (0.56, 2.21) |
| Does child live in a supportive neighborhood |  |  |
| Live in supportive neighborhood | REF |  |
| Do not live in supportive neighborhood | 0.70 | (0.37, 1.34) |
| Parent concerned about child's weight |  |  |
| No, not concerned | REF |  |
| **Yes, concerned it is too high** | **2.97** | **(1.28, 6.87)** |
| Adequate amount of age-appropriate sleep |  |  |
| Child sleeps recommended age-appropriate hours | REF |  |
| Child sleep less than recommended age-appropriate hours | 1.48 | (0.76, 2.89) |
| Child taking medicine for autism |  |  |
| Not taking autism medication | REF |  |
| Taking autism medication | 1.24 | (0.60, 2.60) |
| Parent-rated autism severity |  |  |
| Mild | REF |  |
| Moderate | 1.18 | (0.59, 2.38) |
| Severe | 0.51 | (0.18, 1.45) |

Parent-rated severity of their child’s autism and whether or not their autistic child is taking medicine for their autism were not significant predictors.

## DISCUSSION

Previous studies on correlates of physical activity for autistic children focused only on just one aspect of the child’s life, such as the individual/parental/community influences. This study builds upon previous research into exploring physical activity promotion for autistic children, by using an ecological approach.

Because physical activity, obesity, and autism are interwoven, programs that promote physical activity for autistic children should be flexible to meet the needs of the individual child. In a meta-analysis of physical activity interventions for autistic children, despite the effect size, physical activity interventions had low impact for social functioning or body composition.^16,17^ While current physical activity interventions have effects on muscular strength and endurance, locomotor, manipulative skills, and skill-related fitness, they did not assess physical activities’ holistic impact on the health and well-being of the autistic child, such as improved sleep and weight. In this study, a lack of sleep was associated an aOR of 1.16 for physical inactivity (CI: 1.04-1.30). Additionally, the previous interventions identified in the metanalysis do not utilize efforts like social connectedness efforts like peer-promoters or caregivers to increase physical activity. The study findings showed that when parent concern about their child’s weight was high, their child was more likely to be inactive (aOR = 1.89, CI: 1.59-2.25). Physical activity programs for people with IDD have been successful when incorporating parent caregivers and peer promoters.^18-20^ Future interventions could utilize technology like Fitbit activity watches and scales to track the intervention’s impact on the autistic child’s (with or without co-occurring IDD) sleep, physical activity, and weight by facilitating caregiver and peer involvement.

Additionally, physical education environments had the highest effect size in promoting physical activity.^16^ This finding emphasizes the importance of creating inclusive physical activity environments for autistic children. Inclusive environments allow everyone to take full part in the same program and services by reducing barriers and improving the physical/social well-being.^21^ The Special Olympics and National Center on Health, Physical Activity, and Disability (NCHPAD) have created efforts to promote inclusive physical activity environments, especially at school settings.^21,22^ For example, NCHPAD has a pilot micro-grant to local communities to establish a disability inclusion program to promotion health, physical activity, and weight management.^23^ Disability, physical activity, and governmental organizations could promote ASD inclusion in physical activity programs and activities.

This study is subject to three limitations. First, data from the NSCH is limited to self-reported data. Parents are somewhat likely to provide socially desirable responses for weight and physical activity. This might lead to an underreporting of weight and an overreporting of physical activity. However, the authors expect the social desirability of responses to be the same by ASD/IDD status and thus the aORs would be conservative point estimates. Secondly, the authors cannot determine causality because data from the NSCH are cross-sectional. Finally, interpretations from the study are limited to the variables that are available in the publicly available data set. For example, while the NSCH provides a measure of whether or not the child lives in a metropolitan statistical area, the NSCH does not measure urban versus rural. This is important because these are two different metrics for how the federal government measures density of a community.^24,25^

Having a child with autism, especially with cooccurring IDD, has been associated with many negative health behaviors including physical inactivity and health risks like obesity. Having knowledge about the correlates between autism status and physical inactivity can help organizations, like schools, tailor their programs to be inclusive to autistic children.

## Data Availability

All data is publically available

https://www.childhealthdata.org/

## Notes

**Conflict of Interest:** The authors have no conflicts of interest to declare.

### Competing Interest Statement

The authors have declared no competing interest.

### Funding Statement

This study did not receive any funding

### Author Declarations

Because NSCH data were publicly available, the authors did not seek Institutional Review Board approval.

